# Excess Mortality in Suicide caused by COVID-19 in Japan

**DOI:** 10.1101/2021.02.13.21251670

**Authors:** Junko Kurita, Tamie Sugawara, Yasushi Ohkusa

## Abstract

**Background:** Countermeasures against COVID-19 outbreak such as lockdown and voluntary restrictions against going out adversely affect human stress and economic activity. Particularly, this stress might lead to suicide.

**Object:** We examined excess mortality attributable to suicide caused by COVID-19. Method: We applied the NIID model to suicide deaths from October 2009 through September, 2021 for the whole of Japan by gender. Effects of the great earthquake that struck in eastern Japan on March 11, 2011 were incorporated into the estimation model. Results: Significant excess mortality in suicide was found between July, 2020 and July, 2021 for both genders. However, in August and September, 2021, excess mortality in suicide was detected only in female. It was greater among females than among males. In total, 2950 excess cases of mortality were identified.

**Discussion and Conclusion:** Excess mortality during the four months was more than two times greater than the number of COVID-19 deaths confirmed by PCR testing. Countermeasures against COVID-19 should be chosen carefully in light of suicide effects.

## 1. Introduction

Since the emergence of COVID-19, excess mortality from all causes has been low in Japan [1] before the delta variant strain had emerged. However, we found huge number of excess mortality in August and September, 2021, which were estimated as 4106 and 5854, and were 3.8% and 5.5 % of the baseline. It probably due to the delta variant strain,

However, countermeasures against COVID-19 outbreak such as lockdowns or voluntary restrictions against going out can cause stress and can suppress economic activity. Countermeasures might lower incomes or cause job loss. Such economic stress might lead to suicide [2]. Such difficulties should be counted as part of the costs for countermeasures. Therefore, the present study examined excess mortality attributable to suicide caused by COVID-19.

## 2. Method

The estimation procedure was almost identical to that used for an earlier study [1], except for some points. Excess mortality was defined as the difference between the actual number of deaths and an epidemiological threshold if the actual number of deaths exceeded an epidemiological threshold. The epidemiological threshold is defined as the upper bound of the 95% confidence interval (CI) of the baseline. The baseline is defined as the number of deaths that are likely to have occurred if an influenza outbreak had not occurred. Therefore, if the actual deaths were fewer than the epidemiological threshold, then excess mortality was not inferred. Additionally, we defined negative excess mortality as the difference between the actual number of deaths and the lower bound of 95% CI if the actual deaths were fewer than the lower bound of 95% CI.

Data used for this study were those of monthly deaths from all causes, as reported from 2005 through February, 2021 [3]. The NIID model, the Stochastic Frontier Estimation [4–10], is presented as

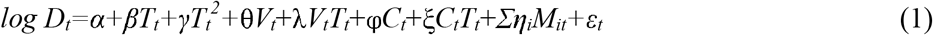

and

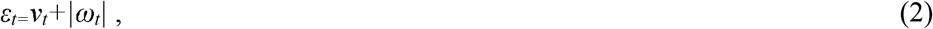

where *D*_*t*_ represents suicide deaths in month/year *t, T*_*t*_ denotes the linear time trend, *V*_*t*_ is a dummy variable for routine pneumococcal vaccination, *C*_*t*_ is a dummy variable for effect of the great earthquake in eastern Japan in March, 2011, and *M*_*it*_ is a dummy variable for the month. Also, *V*_*t*_ takes a value of one after October, 2014; otherwise, it is zero. Similarly, *C*_*t*_ takes a value of one in May, June, or July in 2011; otherwise, it is zero. *M*_*it*_ is one if *t* is the *i-*th month; otherwise, it is zero. Moreover, *ν*_*t*_ and *ω*_*t*_ are stochastic variables as *ν*_*t*_ ∼*N*(*0, μ*^2^) and *ω*_*t*_ ∼*N*(*0,ξ*^2^); they are mutually independent. Although *ν*_*t*_ represents stochastic disturbances, *ω*_*t*_ denotes non-negative deaths attributable to influenza. These disturbance terms in this model are parameterized by two parameters: *ξ*/*μ* and (*μ*^*2*^+*ξ*^*2*^)^*0*.*5*^. If the null hypothesis *ξ*/*μ*=0 is not rejected, then the Stochastic Frontier Estimation model is inappropriate.

The study area encompassed the entire nation of Japan. The study period for estimation was October 2009 through August, 2021 because of data availability. Suicide is defined as X60-X84 in ICD10. We adopted 5% as the level at which significance was inferred for results.

## 3. Results

Figure 1 presents observed suicide deaths, the estimated baseline, and its threshold for males. Figure 2 shows corresponding data for females. These figures showed that males are more numerous than females, but consistently decline is noted in the period for both genders. The sharpest spike occurred around May 2011, probably because of the great earthquake that struck eastern Japan on March 11, 2011. More than two months passed after the event before suicide increased. The second sharpest increase occurred probably in the latest period.

**Figure 1:**
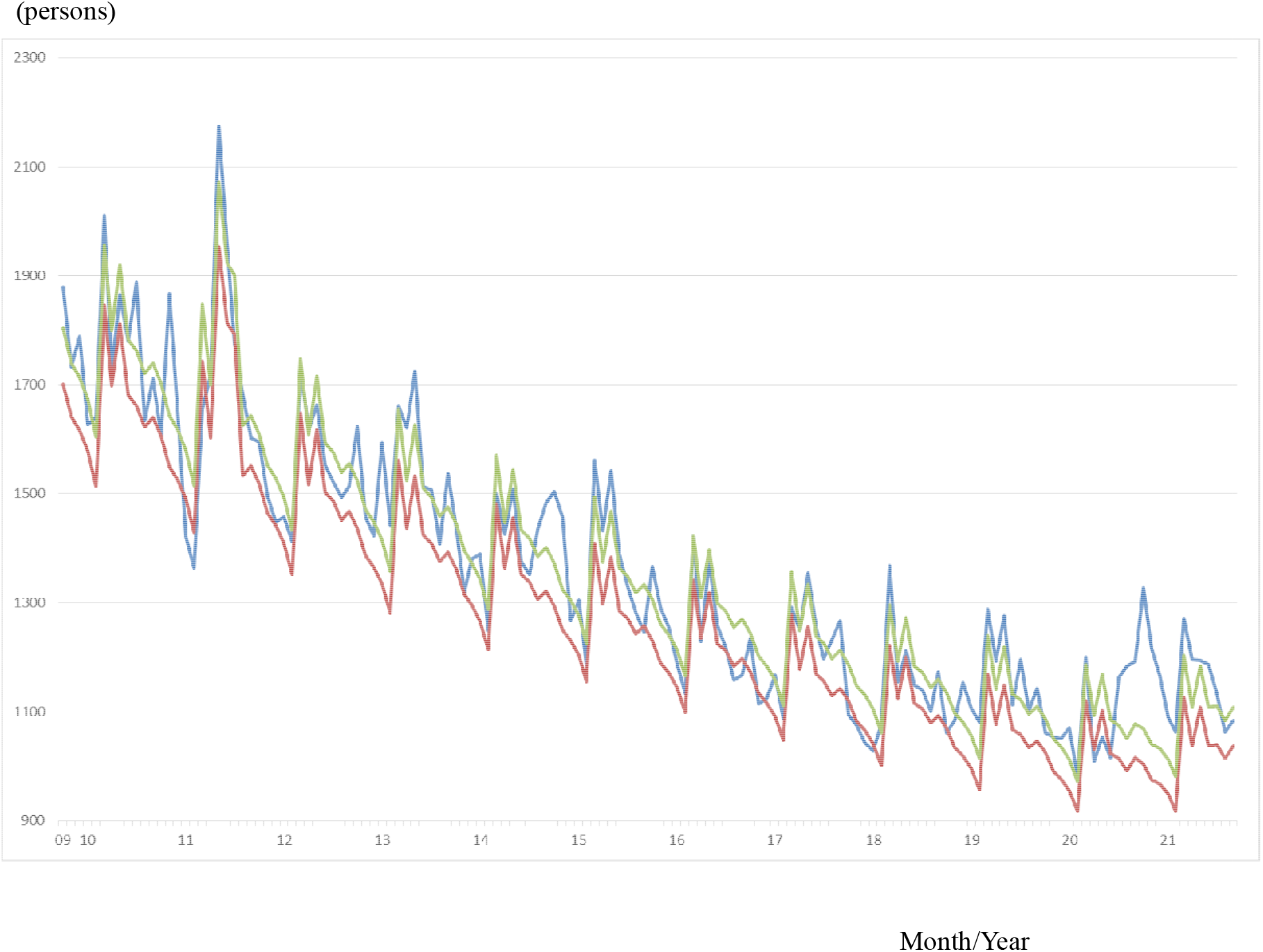
Observations of the estimated baseline and threshold for suicide deaths from October 2009 through September, 2021, among males. Note: The blue line represents observations. The red line represents the estimated baseline. The green line shows its threshold.

**Figure 2:**
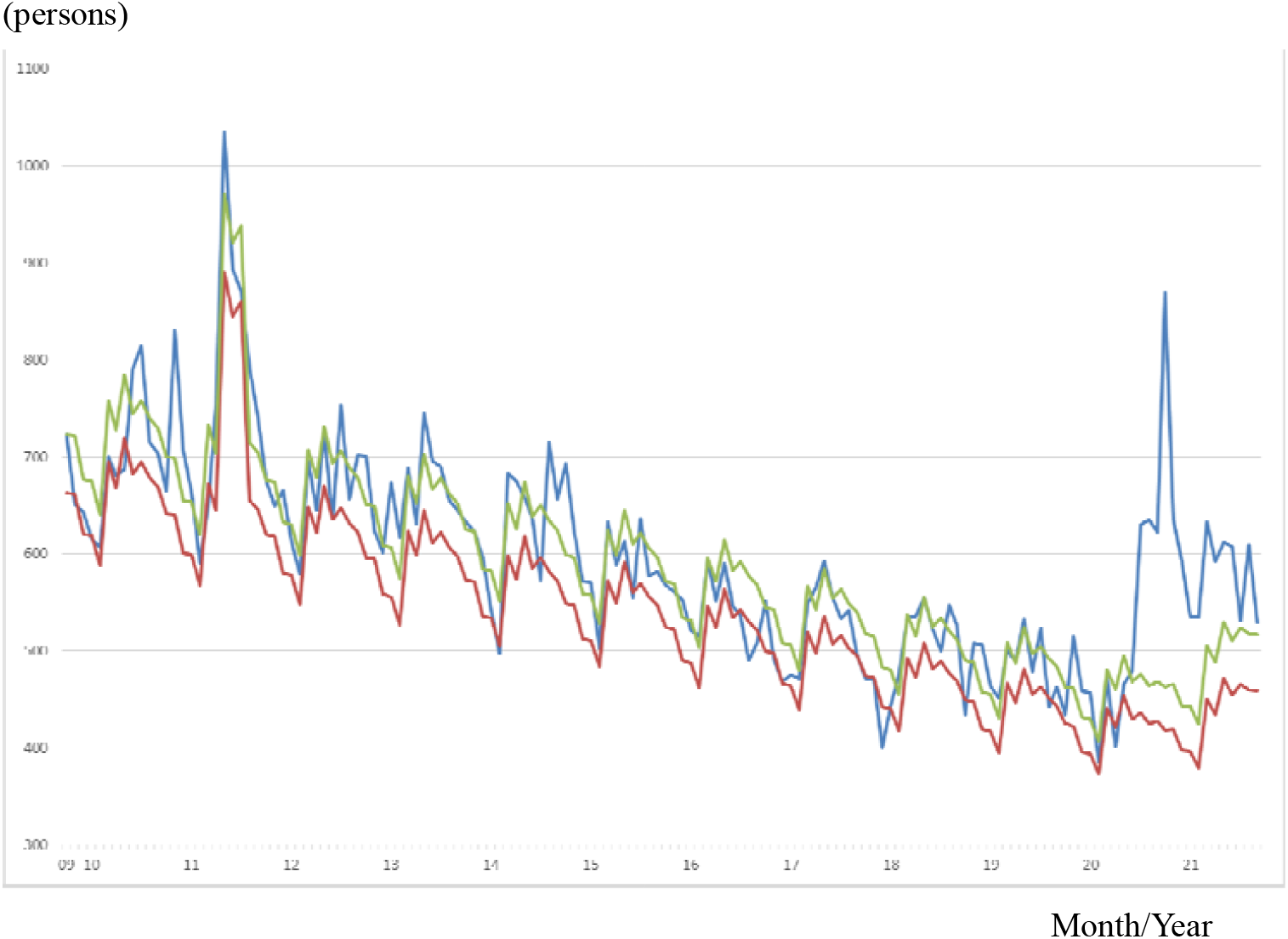
Observations of the estimated baseline and threshold for suicide from October 2009 through September, 2021, among females. Note: The blue line represents observations. The red line represents the estimated baseline. The green line shows its threshold.

Because the estimated coefficients of the cross-term of routine vaccination initiation and time trend, earthquake dummy, and the cross-term of ICD10 revision and time trend were not found to be significant, we dropped these terms from the estimation equation (1). Table 1 presents estimation results without these variables. The estimated coefficients of the great earthquake in eastern Japan were 0.133 for males and 0.247 for females. That result implies that the numbers of suicide deaths jumped respectively by 13% and 25% because of the earthquake. It is noteworthy that the effect on females was greater than on males, although the incidence of suicide death was much greater among males than among females.

**Table 1.** NIID Model estimation results for suicide by gender from October, 2009 through September, 2021 in Japan

| Explanatory variable | Male |  | Female |  |
| --- | --- | --- | --- | --- |
|  | Estimated coefficient | *p*-value | Estimated coefficient | *p*-value |
| Constant | 7.4022 | <0.0004 | 6.4390 | <0.0004 |
| Time trend | -0.004910 | <0.0004 | -0.002571 | 0.001 |
| Time trend <sup>2</sup> | 0.00005637 | 0.114 | 0.000009346 | 0.094 |
| Earthquake | 0.1327 | <0.0004 | 0.2474 | <0.0004 |
| January | -0.01859 | 0.377 | 0.0002747 | 0.993 |
| February | -0.05574 | 0.008 | -0.005012 | 0.140 |
| March | 0.1476 | <0.0004 | 0.1208 | <0.0004 |
| April | 0.06879 | 0.002 | 0.08281 | 0.002 |
| May | 0.1383 | <0.0004 | 0.1605 | <0.0004 |
| June | 0.06852 | 0.016 | 0.1104 | 0.001 |
| July | 0.06223 | 0.011 | 0.1319 | <0.0004 |
| August | 0.04302 | 0.046 | 0.1105 | <0.0004 |
| September | 0.05874 | 0.006 | 0.09846 | 0.002 |
| October | 0.04187 | 0.040 | 0.06022 | 0.025 |
| November | 0.01112 | 0.596 | 0.06072 | 0.026 |
| $\xi/\mu$ | 2.079 | <0.0004 | 2.068 | <0.0004 |
| $(\mu^2 + \xi^2)^{0.5}$ | 0.007422 | <0.0004 | 0.1010 | <0.0004 |
| log likelihood | 213.748 |  | 144.941 |  |

Note: Observations were 146. $\xi^2$ denotes the variance of the non-negative disturbance term. $\mu^2$ represents the variance of the disturbance term. “Earthquake” is a dummy variable for May–July, 2011, which was inferred to be caused by the great earthquake in eastern Japan occurred March 11, 2011.

Figure 3 presents excess mortality attributable to suicide since 2019. Among females, clearly large excess mortality was found since June, 2010. Among males, these were not greater than among females, but excess mortality in male was larger than female in November, 2020. In the 14 months, total excess mortality in suicide was 3246.8.

**Figure 3:**
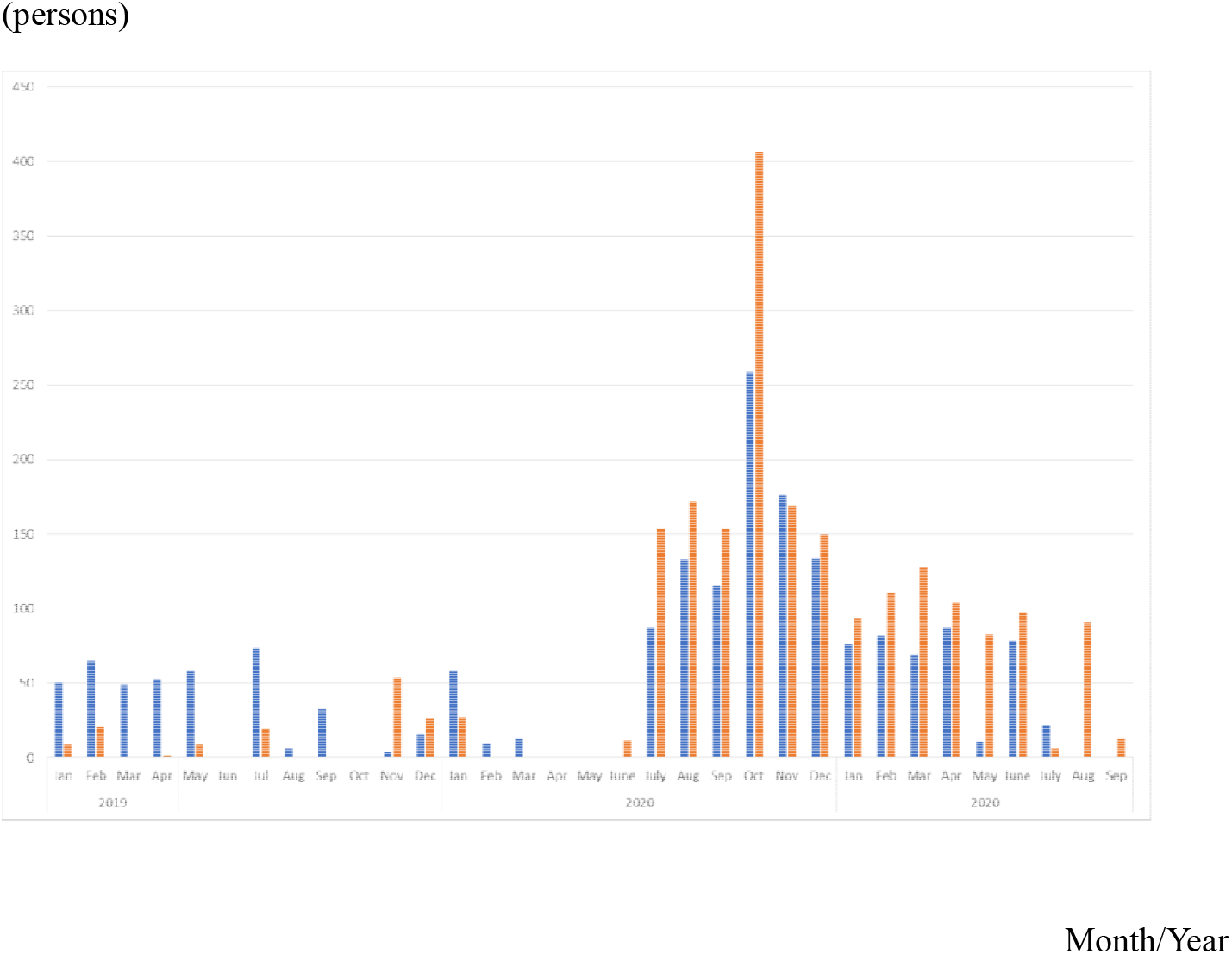
Excess mortality in suicide by gender since 2019 by month in Japan. Note: The blue bars represents excess mortality in suicide among males. Orange bars indicate that among females.

## 4. Discussion

This study applied the NIID model to suicide deaths to detect excess mortality attributable to COVID-19. Results showed that significant excess mortality attributable to suicide was found between July, 2020, and July, 2021, in both gender..

Especially, suicide in females was extremely high in October, 2020. However, in November and December, 2020, excess mortality in suicide decreased. It might be affected by activation of “Go To Travel Campaign” on July 22 which subsidized travel and issuing coupons for shopping at tourist destinations which aims to enforce sightseeing business.

On the other hand, in August and September 2021, excess mortality was not observed in male. This is the first time of no excess morality in male or female since July, 2020. It might indicate that the effect of COVID-19 outbreak might disappear at least in male. Conversely, 91 and 13 suicide excess mortality was observed in female on the same months. This asymmetric phenomena may match the tact that suicide excess mortality in female was much larger than in male. The impact of COVID-19 outbreak for female might continue for a while.

As described earlier, some excess mortality in all causes was found for August and October, 2020, respectively, as 12 and 104 cases. At least 12 cases of excess mortality among all causes in August might result from 304 cases of excess mortality attributable to suicide. At the same time, pneumonia deaths were recorded respectively as 178, 314, and 75 cases of negative excess mortality, in May, June, and July. Those figures were measured as the gap separating the observation and lower bound of 95% CI for baseline if the former was lower than the latter in May, June, and July [11]. In August, negative excess mortality attributable to pneumonia was not found, although the observation was below the baseline. Therefore, excess mortality attributable to suicide was cancelled out partially by negative excess mortality attributable to pneumonia. Therefore, no excess mortality was found for all causes of death. However, in August, negative excess mortality attributable to pneumonia was less than before. Therefore, excess mortality in suicide led to overall excess mortality for all causes, even though it was a very small number. In this sense, excess mortality attributable to suicide might be large in October, although data were not available because excess mortality from all causes was reported as a larger number than in August.

Some researchers in Japan have emphasized considerable excess mortality from all causes of death through June of around 48 thousand at maximum [12] when using the Farrington algorithm [13] and EuroMOMO [14], which was three times larger than the number of death confirmed by PCR test. This study measured excess mortality as the gap separating observations and baseline, not a threshold, as in prefectures where observations were higher than the threshold Therefore, their estimated too huge excess mortality might seriously mislead people into adopting greater risks of infection by COVID-19 among the general population.

To ascertain the most appropriate countermeasures for COVID-19 in Japan, cost-effectiveness analysis is necessary. At that time, loss of quality of life in should be counted among the costs of restriction of economic activity as a major part of countermeasures. It remains as a subject for our next research challenge.

## 5. Conclusion

The obtained results show excess mortality in suicide between July, 2020 and July, 2021 in both of gender. Especially, suicide in female in October, 2020 was remarkable. Moreover, in August and September, 2021, the suicide excess mortality was observed only in female. Though the number of suicide was larger in male than female, impact of COVID-19 outbreak for female may be larger than male. Continued careful monitoring of excess mortality attributable to suicide is expected to be necessary so as to control counter measure for COVID-19 not to be too restrictive for business and daily life.

The present study is based on the authors’ opinions: it does not reflect any stance or policy of their professionally affiliated bodies.

## Data Availability

National Institute of Infectious Diseases, Excess mortality in Japan, on October 2020.

https://www.niid.go.jp/niid/ja/from-idsc/493-guidelines/10150-excess-mortality-21jan.html

## 6. Acknowledgement

We acknowledge Dr. Nobuhiko Okabe, Kawasaki City Institute for Public Health, Dr.Kiyosu Taniguchi, National Hospital Organization Mie National Hospital, and Dr.Nahoko Shindo, WHO for their helpful support.

## 8. Ethical considerations

All information used for this study was published on the web site of MHLW [12]. Therefore, no ethical issue is presented.

